# Association of Age With Guideline-Directed Medical Therapy and Left Ventricular Reverse Remodeling

**DOI:** 10.64898/2026.09.04.26362221

**Authors:** In-Chang Hwang, Jiesuck Park, Nan Young Bae, Jaehyun Lim, Soongu Kwak, Minjung Bak, Hong-Mi Choi, Jun-Bean Park, Yeonyee E. Yoon, Seung-Pyo Lee, Yong-Jin Kim, Goo-Yeong Cho, Hyung-Kwan Kim

## Abstract

**Background:** Comprehensive guideline-directed medical therapy (GDMT) remains underused in older patients with HFrEF, but whether the potential for treatment-associated myocardial recovery is preserved with advancing age remains uncertain.

**Objectives:** To characterize age-related GDMT gaps and determine whether GDMT intensity is associated with left ventricular (LV) reverse remodeling across age and subsequent outcomes.

**Methods:** We studied 1,795 patients with HFrEF receiving angiotensin receptor-neprilysin inhibitor (ARNI)-based therapy in the multicenter STRATS-HF-ARNI registry. GDMT intensity was defined by the number of foundational therapies used during the first year after ARNI initiation. Serial LV remodeling and subsequent death or HF hospitalization were evaluated using multivariable models and a 1-year landmark analysis.

**Results:** Triple/quadruple GDMT was achieved in 59.1%, 50.8%, 53.4%, and 43.0% of patients aged <60, 60–69, 70–79, and ≥80 years, respectively (P<0.001). Older age and renal dysfunction independently predicted less comprehensive GDMT. Each additional GDMT pillar was associated with greater improvements in LVEF (+1.23 percentage-points; P=0.012) and LV global longitudinal strain (+0.32 percentage-points; P=0.049) and greater reductions in LV volumes. Substantial remodeling remained evident at advanced age. Greater LV reverse remodeling predicted lower subsequent risk, and exploratory bootstrap analyses supported significant indirect associations between greater GDMT intensity and lower risk through all four LV remodeling measures.

**Conclusions:** Older patients received less comprehensive GDMT despite preserved potential for LV reverse remodeling. Greater GDMT intensity was associated with greater myocardial recovery, which may represent a pathway to better prognosis. These findings support intensive GDMT implementation in older patients when clinically tolerated.

## Introduction

Contemporary guideline-directed medical therapy (GDMT) for heart failure with reduced ejection fraction (HFrEF) comprises four foundational therapies: angiotensin receptor-neprilysin inhibitor (ARNI) or angiotensin-converting enzyme inhibitor/angiotensin receptor blocker, beta-blocker (BB), mineralocorticoid receptor antagonist (MRA), and sodium-glucose cotransporter-2 inhibitor (SGLT2i).^1–3^ Recent studies and recommendations emphasize early implementation of these complementary therapies rather than prolonged sequential titration.^3,4^ This approach may be particularly challenging in older patients, in whom multimorbidity and concerns regarding treatment tolerance may limit multidrug therapy.

Despite these recommendations, comprehensive GDMT remains underused, particularly in older patients.^5–9^ Advancing age is accompanied by renal dysfunction, hypotension, hyperkalemia, bradycardia, multimorbidity, and frailty, which may limit treatment intensification.^5,6,9^ Nevertheless, randomized-trial subgroup analyses show preserved efficacy of individual foundational therapies across age.^10–13^ Thus, a gap persists between the expected benefits of GDMT and its implementation in older patients.

Contemporary HF therapies promote left ventricular (LV) reverse remodeling, including improvement in LV ejection fraction (LVEF), LV volumes, and LV global longitudinal strain (LVGLS).^14–20^ Prior studies have demonstrated reverse remodeling in older patients but have largely focused on individual therapies, whereas studies of age-related GDMT underuse have rarely examined remodeling according to overall treatment intensity. Whether greater multidrug GDMT intensity is associated with greater LV recovery across the age spectrum therefore remains incompletely characterized. We assessed age-related differences in comprehensive and four-pillar GDMT, factors associated with incomplete implementation, and the association of GDMT intensity with LV reverse remodeling across age, as well as subsequent clinical outcomes.

## Methods

### Study population

This study used the multicenter STrain for Risk Assessment and Therapeutic Strategies in patients with Heart Failure treated with Angiotensin Receptor-Neprilysin Inhibitor (STRATS-HF-ARNI) registry of 2,757 consecutive patients with HFrEF treated with ARNI at Seoul National University Hospital and Seoul National University Bundang Hospital from 2017 through 2022.^15,17,20,21^ The registry was registered with the Clinical Research Information Service (KCT0008098). Institutional Review Boards at both centers approved the study and waived individual informed consent because of its retrospective observational design.

Patients were eligible if they were aged ≥20 years, had baseline echocardiography with LVEF measurement, and maintained ARNI for ≥6 months. Patients without baseline echocardiography, those who discontinued ARNI within 6 months, those aged <20 years, and those with insufficient baseline echocardiographic data were excluded. GDMT analyses used the overall eligible cohort; reverse-remodeling analyses used patients with baseline and approximately 1-year follow-up echocardiography.

### Age, Calendar Period, and GDMT Intensity

Patients were categorized as <60, 60-69, 70-79, or ≥80 years. The four foundational classes were ARNI, BB, MRA, and SGLT2i. Because all patients received ARNI, GDMT intensity was determined by concomitant BB, MRA, and SGLT2i use during the first year after ARNI initiation and classified as mono-, dual-, triple-, or quadruple therapy. Comprehensive GDMT was defined as triple/quadruple therapy; quadruple GDMT required all four classes. To account for temporal changes in clinical evidence, guideline recommendations, and adoption of contemporary HFrEF therapies in clinical practice during the study period, patients were additionally categorized according to the year of ARNI initiation into calendar periods of 2017-2019, 2020, 2021, and 2022. Medication exposure was defined according to treatment achieved during the first year after ARNI initiation rather than medication status at a single baseline visit, thereby reflecting treatment implementation during the interval in which LV reverse remodeling was assessed.

### Factors Associated With GDMT Implementation

Potential correlates included renal dysfunction (estimated glomerular filtration rate [eGFR] <60 mL/min/1.73 m²), hyperkalemia (potassium ≥5.0 mmol/L), hypotension (systolic blood pressure <100 mmHg), and bradycardia (heart rate <60 beats/min). Bradycardia was not included in multivariable models because heart rate was unavailable in a substantial proportion. Factors associated with failure to achieve comprehensive and quadruple GDMT were evaluated using multivariable logistic regression as described below.

### Echocardiographic Evaluation and Assessment of Reverse Remodeling

Transthoracic echocardiography was performed according to contemporary recommendations.^22^ LVEF, LV end-diastolic volume (LVEDV), and LV end-systolic volume (LVESV) were measured by biplane Simpson’s methods. LVGLS was measured by 2-dimensional speckle tracking and expressed as absolute magnitude.^19,23,24^ Changes were calculated as follow-up minus baseline; positive ΔLVEF and ΔLVGLS and negative ΔLVEDV and ΔLVESV represented favorable remodeling.

Changes in LVEF, LVGLS, LVEDV, and LVESV were compared across GDMT categories, and ordinal trends were assessed. Associations of GDMT intensity with remodeling were evaluated using multivariable linear regression as described below.

### Study Outcomes

The treatment implementation outcomes were failure to achieve comprehensive GDMT and failure to achieve quadruple GDMT. The principal echocardiographic outcomes were changes in LVEF, LVGLS, LVEDV, and LVESV. Clinical outcomes included all-cause mortality, HF hospitalization, and their composite. Because GDMT intensity was assessed during the first year after ARNI initiation, clinical outcomes were evaluated using a 1-year landmark analysis, excluding patients with events before or without follow-up to the landmark. Among patients with available follow-up echocardiography, the associations of changes in LVEF, LVGLS, LVEDV, and LVESV with the subsequent composite of all-cause death or HF hospitalization were additionally evaluated using the same landmark framework.

## Statistical Analysis

Continuous variables are presented as mean±standard deviation or median with interquartile range, as appropriate, and categorical variables as frequency with percentage. Comparisons across age groups were performed using analysis of variance or the Kruskal-Wallis test for continuous variables and the chi-square test for categorical variables. GDMT use was compared across age groups and calendar periods. Multivariable logistic regression was used to identify factors associated with failure to achieve comprehensive and quadruple GDMT. Models included age, sex, eGFR, diabetes mellitus, atrial fibrillation, coronary artery disease, baseline LVEF, and calendar year; age was modeled per 10-year increase. An additional analysis of comprehensive GDMT was performed among patients aged ≥70 years.

In the echocardiographic subgroup, multivariable linear regression evaluated GDMT intensity per additional foundational therapy in relation to changes in LVEF, LVGLS, LVEDV, and LVESV. Models included age, sex, body mass index, systolic blood pressure, eGFR, diabetes mellitus, atrial fibrillation, coronary artery disease, and the corresponding baseline value. Age-stratified analyses were adjusted for age, sex, systolic blood pressure, diabetes mellitus, coronary artery disease, and the corresponding baseline value; age-by-GDMT interactions were tested.

For clinical outcomes, Kaplan-Meier analyses and Cox proportional-hazards regression were performed using a 1-year landmark approach. GDMT intensity was examined both per additional foundational therapy and categorically as mono-, dual-, triple-, or quadruple therapy. The multivariable composite-outcome model included age, sex, systolic blood pressure, eGFR, diabetes mellitus, atrial fibrillation, coronary artery disease, baseline LVEF, and GDMT intensity. Among patients with serial echocardiography, separate Cox models evaluated the associations of changes in LVEF, LVGLS, LVEDV, and LVESV with the subsequent composite outcome because of intercorrelation among remodeling measures. Models were adjusted for clinical covariates and the corresponding baseline echocardiographic parameter. Effect estimates were expressed per 5-percentage-point improvement in LVEF, 1-percentage-point improvement in absolute LVGLS, and 10-mL reduction in LVEDV or LVESV.

Exploratory indirect-effect analyses evaluated each LV remodeling measure as a potential intermediate pathway between GDMT intensity and the subsequent composite outcome. Linear and Cox regression modeled the GDMT-remodeling and remodeling-outcome associations, respectively. Indirect effects were calculated by the product-of-coefficients method on the log-hazard scale and expressed as indirect HRs. Percentile 95% CIs were derived from 1,000 bootstrap resamples with both models refitted in each sample. These analyses were not intended to establish causal mediation.

All statistical tests were two-sided, and P values <0.05 were considered statistically significant.

## Results

### Baseline Characteristics According to Age

Among 1,795 patients, 582 (32.4%) were aged <60 years, 463 (25.8%) 60-69 years, 487 (27.1%) 70-79 years, and 263 (14.7%) ≥80 years (**Table 1**). With advancing age, chronic kidney disease, coronary artery disease, and atrial fibrillation were more frequent, and median eGFR declined from 89.6 to 56.3 mL/min/1.73 m² (P<0.001). Older patients also had lower body mass index and progressively higher NT-proBNP concentrations. Baseline LV volumes were smaller with advancing age, whereas left atrial volume index and E/e′ ratio were greater. Use of BB decreased from 92.8% in patients aged <60 years to 87.5% in those aged ≥80 years, and MRA use declined from 55.0% to 40.7%, whereas SGLT2i use was numerically lower but did not differ significantly across age groups.

**Table 1.** Baseline Characteristics According to Age Group.

| Variable | <60 years<br>(n = 582) | 60–69 years<br>(n = 463) | 70–79 years<br>(n = 487) | ≥80 years<br>(n = 263) | P value |
| --- | --- | --- | --- | --- | --- |
| Age, year | 51.1 (43.8–56.5) | 65.0 (62.4–67.5) | 75.0 (73.0–77.1) | 83.0 (81.6–86.0) | <0.001 |
| Male | 458 (78.7) | 337 (72.8) | 297 (61.0) | 145 (55.1) | <0.001 |
| BMI, kg/m<sup>2</sup> | 25.3 (22.7–28.4) | 24.4 (22.5–26.4) | 24.0 (21.8–25.9) | 23.4 (20.9–25.8) | <0.001 |
| Systolic blood pressure, mmHg | 118 (107–132) | 120 (106–135) | 121 (109–135) | 121 (110–135) | 0.367 |
| Heart rate, beats/min | 75 (69–87) | 75 (67–88) | 72 (65–81) | 72 (64–79) | 0.003 |
| **Comorbidities** |  |  |  |  |  |
| Hypertension | 139 (23.9) | 166 (35.9) | 212 (43.6) | 114 (43.3) | <0.001 |
| Diabetes mellitus | 130 (22.3) | 161 (34.8) | 179 (36.8) | 68 (25.9) | <0.001 |
| Chronic kidney disease | 69 (11.9) | 95 (20.5) | 150 (30.8) | 128 (48.7) | <0.001 |
| Coronary artery disease | 157 (27.0) | 187 (40.4) | 184 (37.8) | 111 (42.2) | <0.001 |
| Atrial fibrillation | 101 (17.4) | 118 (25.5) | 162 (33.3) | 72 (27.4) | <0.001 |
| **HF medications** |  |  |  |  |  |
| Beta-blocker | 540 (92.8) | 416 (89.8) | 429 (88.1) | 230 (87.5) | 0.031 |
| MRA | 320 (55.0) | 215 (46.4) | 248 (50.9) | 107 (40.7) | <0.001 |
| SGLT2 inhibitor | 127 (21.8) | 96 (20.7) | 93 (19.1) | 42 (16.0) | 0.232 |
| **Laboratory findings** |  |  |  |  |  |
| BUN, mg/dL | 17 (14–22) | 20 (15–26) | 21 (17–28) | 23 (18–31) | <0.001 |
| Creatinine, mg/dL | 0.94 (0.80–1.17) | 0.99 (0.83–1.30) | 1.06 (0.84–1.35) | 1.09 (0.89–1.38) | <0.001 |
| eGFR, mL/min/1.73 m<sup>2</sup> | 89.6 (69.9–103.1) | 75.9 (54.2–90.8) | 63.6 (47.9–80.2) | 56.3 (39.9–70.9) | <0.001 |
| Hemoglobin, g/dL | 14.4 (12.9–15.8) | 13.3 (12.0–14.6) | 12.7 (11.3–14.0) | 12.2 (11.1–13.6) | <0.001 |
| Sodium, mmol/L | 140 (138–141) | 140 (138–141) | 140 (138–142) | 140 (138–142) | 0.237 |
| Potassium, mmol/L | 4.3 (4.1–4.6) | 4.4 (4.1–4.7) | 4.4 (4.1–4.8) | 4.4 (4.0–4.8) | 0.250 |
| Total cholesterol, mg/dL | 161 (133–197) | 144 (120–179) | 142 (119–174) | 138 (117–171) | <0.001 |
| NT-proBNP, pg/mL | 940 (320–2674) | 1353 (500–3427) | 2319 (938–4808) | 3405 (1372–7824) | <0.001 |
| **Baseline echocardiography** |  |  |  |  |  |
| LV end-diastolic dimension, mm | 61 (55–65) | 60 (55–64) | 58 (53–63) | 56 (52–61) | <0.001 |
| LV end-systolic dimension, mm | 50 (45–56) | 49 (44–54) | 48 (42–53) | 46 (42–51) | <0.001 |
| LV end-diastolic volume, mL | 173 (137–218) | 159 (127–199) | 143 (113–186) | 132 (104–163) | <0.001 |
| LV end-systolic volume, mL | 123 (90–165) | 109 (83–144) | 95 (74–130) | 92 (71–116) | <0.001 |
| LV ejection fraction, % | 30.0 (23.0–35.0) | 31.9 (25.3–36.0) | 32.2 (27.1–36.0) | 31.1 (26.0–35.2) | <0.001 |
| LV mass index, g/m<sup>2</sup> | 131.0 (108.8–159.5) | 137.2 (112.4–161.9) | 135.9 (112.2–157.6) | 140.4 (115.6–158.9) | 0.340 |
| LA volume index, mL/m<sup>2</sup> | 50.3 (37.5–64.6) | 54.0 (41.4–72.6) | 61.5 (46.2–80.2) | 62.2 (46.1–77.6) | <0.001 |
| Mitral E velocity, m/s | 0.75 (0.56–0.97) | 0.70 (0.50–0.93) | 0.79 (0.51–1.04) | 0.76 (0.53–0.97) | 0.036 |
| e' velocity, cm/s | 5.2 (4.0–6.4) | 4.5 (3.7–5.4) | 4.2 (3.1–5.3) | 3.9 (3.0–5.0) | <0.001 |
| E/e' ratio | 13.8 (10.0–20.1) | 15.0 (11.0–21.1) | 17.0 (12.6–24.2) | 18.1 (13.4–24.2) | <0.001 |
| PASP, mmHg | 32.0 (26.2–43.4) | 34.0 (26.2–46.0) | 34.2 (28.0–46.0) | 36.4 (28.0–46.0) | 0.016 |
| LV global longitudinal strain, %* | 9.3 (7.0–11.7) | 9.5 (7.8–12.1) | 9.8 (7.8–12.0) | 8.6 (6.9–11.0) | 0.002 |
| LA reservoir strain, %* | 13.3 (8.9–19.0) | 13.2 (8.9–18.8) | 12.5 (8.3–16.9) | 11.9 (8.2–16.7) | <0.001 |

Data are presented as median (interquartile range) for continuous variables and n (%) for categorical variables. P values were obtained using the Kruskal-Wallis test for continuous variables and chi-square test for categorical variables.
\*LV global longitudinal strain and LA reservoir strain are presented as absolute values.
Abbreviations: BMI, body mass index; eGFR, estimated glomerular filtration rate; HF, heart failure; LA, left atrial; LV, left ventricular; MRA, mineralocorticoid receptor antagonist; NT-proBNP, N-terminal pro-B-type natriuretic peptide; PASP, pulmonary artery systolic pressure; SGLT2, sodium-glucose cotransporter-2.

Comprehensive (triple or quadruple) GDMT was achieved in 59.1%, 50.8%, 53.4%, and 43.0% across increasing age groups (P<0.001), and quadruple therapy in 15.1%, 10.8%, 10.7%, and 8.0% (P=0.012) (**Figure 1**). Age-related differences in comprehensive and quadruple GDMT were generally observed during the earlier calendar periods but were attenuated in 2022. Quadruple therapy increased across calendar periods, reaching 28.3%, 17.6%, 34.6%, and 18.5% in the <60, 60-69, 70-79, and ≥80-year groups, respectively, in 2022. Among patients receiving dual therapy, ARNI+BB predominated (672/749, 89.7%); among those receiving triple therapy, ARNI+BB+MRA predominated (610/740, 82.4%) (**Supplementary Table S1**).

**Figure 1.**
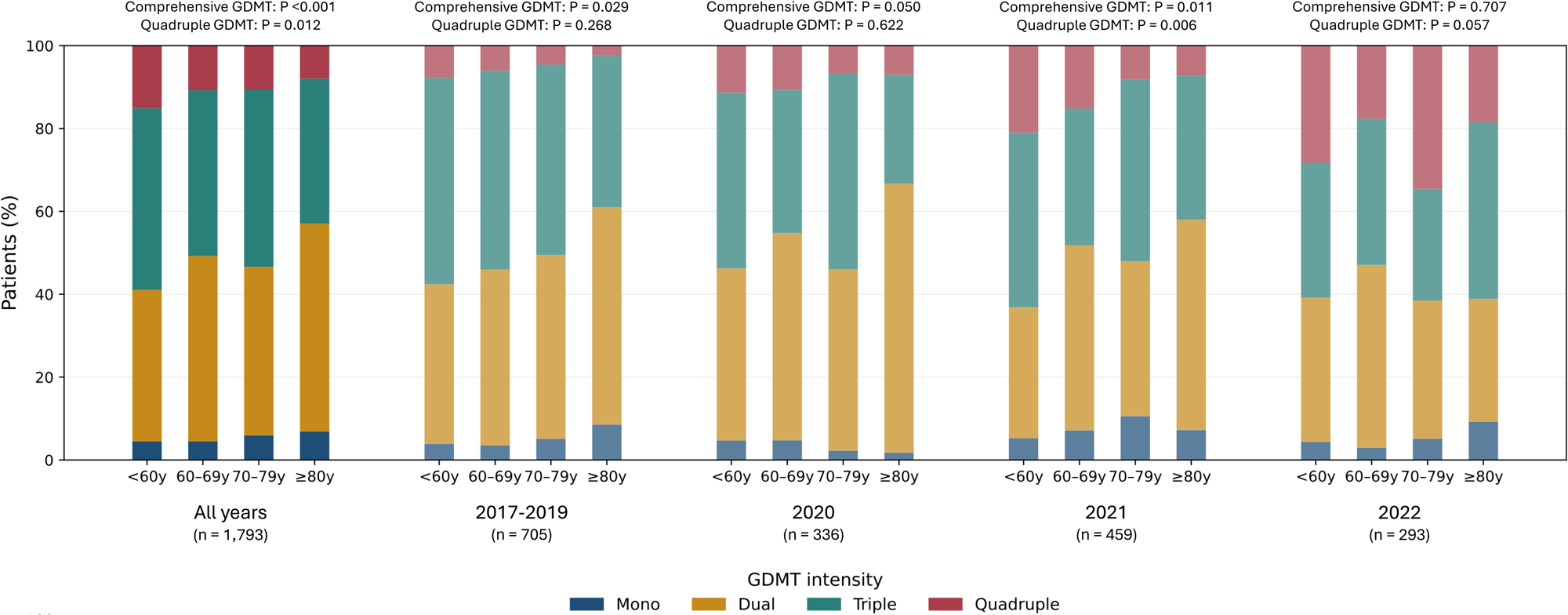
Temporal Changes in Guideline-Directed Medical Therapy According to Age. Distribution of mono-, dual-, triple-, and quadruple GDMT by age group and calendar period of ARNI initiation. P values compare comprehensive (triple/quadruple) and quadruple GDMT across age groups. Calendar periods were 2017–2019, 2020, 2021, and 2022. ARNI, angiotensin receptor-neprilysin inhibitor; GDMT, guideline-directed medical therapy.

### Factors Associated With GDMT Implementation

Advancing age independently predicted failure to achieve comprehensive GDMT (adjusted OR per 10 years, 1.09; 95% CI, 1.01-1.18; P=0.036) and quadruple GDMT (adjusted OR, 1.19; 95% CI, 1.06-1.35; P=0.005) (**Supplementary Table S2**). eGFR <60 mL/min/1.73 m² was associated with higher odds of failure at both thresholds (adjusted OR, 1.88 [95% CI, 1.49-2.36] and 2.26 [95% CI, 1.53-3.33], respectively; both P<0.001). Diabetes mellitus was associated with lower odds of failure at both treatment thresholds. More recent calendar year was also associated with lower odds of failure to achieve comprehensive and quadruple GDMT (adjusted OR per year, 0.88 [95% CI, 0.82-0.94] and 0.55 [95% CI, 0.48-0.63], respectively; both P<0.001). Coronary artery disease was modestly associated with higher odds of failure to achieve comprehensive GDMT (adjusted OR, 1.27; 95% CI, 1.01-1.58; P=0.037).

Among patients aged ≥70 years, advancing age and eGFR <60 mL/min/1.73 m² were associated with higher odds of failure to achieve comprehensive GDMT, whereas diabetes mellitus was associated with lower odds of failure (**Supplementary Table S3**). More recent calendar year was also associated with lower odds of failure to achieve comprehensive GDMT (adjusted OR per year, 0.82; 95% CI, 0.74-0.91; P<0.001).

### LV Reverse Remodeling According to Age and GDMT Intensity

Follow-up echocardiography was available in 1,194 patients: 395 (67.9%), 327 (70.6%), 315 (64.7%), and 147 (55.9%) across increasing age groups. LV reverse remodeling was more prominent with more intense GDMT: mean ΔLVEF was +6.1%, +9.7%, +11.4%, and +12.1% across mono-, dual-, triple-, and quadruple therapy; ΔLVGLS was +1.83%, +1.96%, +2.33%, and +3.08%; ΔLVEDV was-17.8,-29.0,-31.6, and-34.3 mL; and ΔLVESV was-20.0,-31.9,-34.7, and-39.5 mL, respectively (**Figure 2**). Although age-stratified associations were strongest in patients aged <60 years, favorable remodeling remained evident in older groups. Among patients aged ≥80 years, mean ΔLVEF was +4.7%, +10.3%, +10.7%, and +11.8% across mono-, dual-, triple-, and quadruple therapy, respectively; corresponding improvements in LVGLS were +0.09%, +1.80%, +1.94%, and +2.54%.

**Figure 2.**
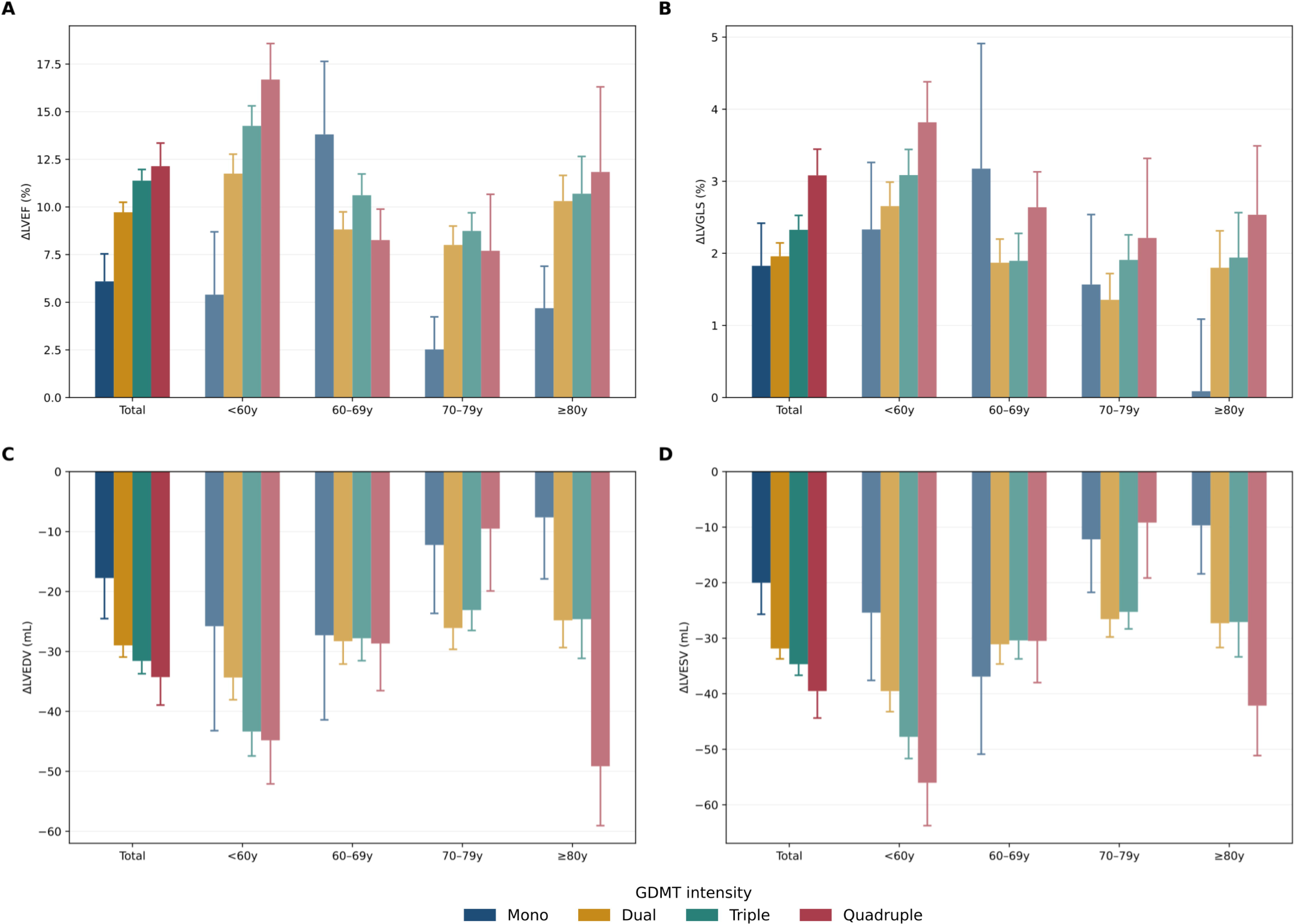
Left Ventricular Reverse Remodeling According to Age and GDMT Intensity. Changes in (A) LVEF, (B) absolute LVGLS, (C) LVEDV, and (D) LVESV from baseline to approximately 1 year according to age and GDMT intensity. Bars show mean±SE; P values indicate adjusted trends within age groups. Positive LVEF/LVGLS and negative LVEDV/LVESV changes indicate favorable remodeling. GDMT, guideline-directed medical therapy; LVEDV, left ventricular end-diastolic volume; LVEF, left ventricular ejection fraction; LVESV, left ventricular end-systolic volume; LVGLS, left ventricular global longitudinal strain.

After adjustment, each additional GDMT pillar was independently associated with greater LVEF improvement (β=+1.23 percentage points; 95% CI, 0.27-2.18; P=0.012), LVGLS improvement (β=+0.32%; 95% CI, 0.00 to 0.63; P=0.049), LVEDV reduction (β=+4.69 mL; 95% CI, 1.12-8.26; P=0.010), and LVESV reduction (β=+4.50 mL; 95% CI, 1.19-7.82; P=0.008) (**Table 2**).

**Table 2.** Multivariable Predictors of Left Ventricular Reverse Remodeling in the Echocardiographic Subgroup.

| Variable | LVEF improvement |  | LVGLS improvement |  | LVEDV reduction |  | LVESV reduction |  |
| --- | --- | --- | --- | --- | --- | --- | --- | --- |
| | $\beta$ (95% CI) | P value | $\beta$ (95% CI) | P value | $\beta$ (95% CI) | P value | $\beta$ (95% CI) | P value |
| Age, per 10-year increase | -0.94 (-1.50 to -0.37) | 0.001 | -0.42 (-0.60 to -0.23) | <0.001 | -1.38 (-3.54 to 0.77) | 0.209 | -2.57 (-4.58 to -0.57) | 0.012 |
| Male | -3.87 (-5.35 to -2.39) | <0.001 | -1.08 (-1.57 to -0.60) | <0.001 | -10.98 (-16.85 to -5.11) | <0.001 | -10.36 (-15.75 to -4.97) | <0.001 |
| BMI, per 5 kg/m<sup>2</sup> increase | -0.32 (-0.95 to 0.31) | 0.321 | -0.38 (-0.58 to -0.17) | <0.001 | -1.45 (-3.83 to 0.92) | 0.230 | -1.29 (-3.48 to 0.91) | 0.251 |
| SBP, per 10-mmHg increase | 0.65 (0.30 to 0.99) | <0.001 | 0.11 (-0.00 to 0.22) | 0.061 | 1.20 (-0.09 to 2.50) | 0.069 | 1.37 (0.17 to 2.57) | 0.025 |
| eGFR, per 10 mL/min/1.73 m<sup>2</sup> increase | 0.11 (-0.17 to 0.40) | 0.435 | -0.01 (-0.10 to 0.09) | 0.899 | 0.28 (-0.80 to 1.35) | 0.614 | 0.37 (-0.63 to 1.37) | 0.467 |
| Diabetes mellitus | -1.42 (-3.00 to 0.16) | 0.078 | -0.74 (-1.26 to -0.22) | 0.005 | -9.22 (-15.17 to -3.27) | 0.002 | -8.87 (-14.39 to -3.34) | 0.002 |
| Atrial fibrillation | 1.30 (-0.29 to 2.90) | 0.110 | -0.07 (-0.59 to 0.45) | 0.789 | 1.17 (-4.94 to 7.27) | 0.708 | -0.07 (-5.73 to 5.59) | 0.980 |
| Coronary artery disease | -4.10 (-5.56 to -2.65) | <0.001 | -1.10 (-1.58 to -0.63) | <0.001 | -9.98 (-15.42 to -4.54) | <0.001 | -11.93 (-16.98 to -6.87) | <0.001 |
| GDMT intensity, per additional pillar | 1.23 (0.27 to 2.18) | 0.012 | 0.32 (0.00 to 0.63) | 0.049 | 4.69 (1.12 to 8.26) | 0.010 | 4.50 (1.19 to 7.82) | 0.008 |
| Baseline LVEF, per 10% increase | -6.50 (-7.34 to -5.66) | <0.001 | — | — | — | — | — | — |
| Baseline LVGLS, per 5% increase | — | — | -3.19 (-3.52 to -2.86) | <0.001 | — | — | — | — |
| Baseline LVEDV, per 10-mL increase | — | — | — | — | 3.28 (2.83 to 3.72) | <0.001 | — | — |
| Baseline LVESV, per 10-mL increase | — | — | — | — | — | — | 3.92 (3.44 to 4.41) | <0.001 |

Positive $\beta$ values indicate greater improvement in LVEF or LVGLS and greater reduction in LVEDV or LVESV. Each model included age, sex, BMI, systolic blood pressure, estimated glomerular filtration rate, diabetes mellitus, atrial fibrillation, coronary artery disease, GDMT intensity, and the corresponding baseline echocardiographic value.
Abbreviations: BMI, body mass index; CI, confidence interval; eGFR, estimated glomerular filtration rate; GDMT, guideline-directed medical therapy; LV, left ventricular; LVEDV, left ventricular end-diastolic volume; LVEF, left ventricular ejection fraction; LVESV, left ventricular end-systolic volume; LVGLS, left ventricular global longitudinal strain; SBP, systolic blood pressure.

### Clinical Outcomes and Exploratory Prognostic Analyses

In 1-year landmark analyses, event risk increased with age (**Supplementary Figure S1**). Compared with monotherapy, dual and quadruple therapy were associated with lower all-cause mortality, but no consistent stepwise association across GDMT intensity was observed for HF hospitalization or the composite outcome (**Supplementary Table S4**). The composite outcome according to GDMT intensity in the overall population and across age groups is shown in **Figure 3**. In the composite-outcome predictor model, older age predicted higher risk (adjusted HR per 10 years, 1.37; 95% CI, 1.21-1.56; P<0.001), whereas higher eGFR predicted lower risk (adjusted HR per 10 mL/min/1.73 m², 0.87; 95% CI, 0.82-0.93; P<0.001) (**Supplementary Table S5**). GDMT intensity, analyzed per additional pillar, was not independently associated with the composite outcome (adjusted HR, 0.94; 95% CI, 0.76-1.16; P=0.545). Kaplan-Meier curves for all-cause mortality and HF hospitalization according to GDMT intensity are shown in **Supplementary Figures S2** and **S3**, respectively.

**Figure 3.**
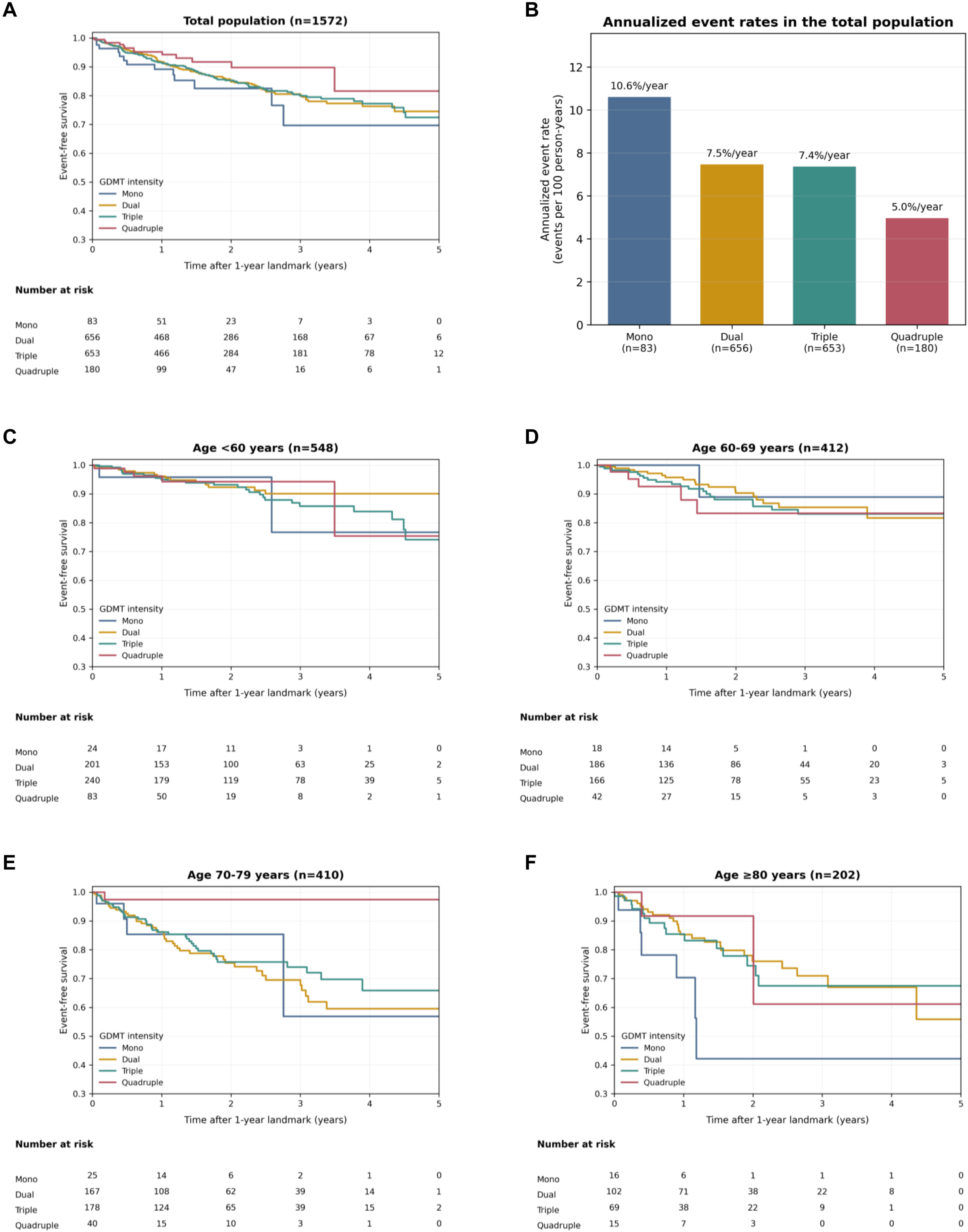
Composite Clinical Outcome According to GDMT Intensity and Age. One-year landmark analysis of the composite of all-cause death or HF hospitalization according to GDMT intensity. (A) Kaplan-Meier curves in the overall population. (B) Annualized composite event rates according to GDMT intensity in the overall population. (C–F) Kaplan-Meier curves in patients aged <60, 60–69, 70–79, and ≥80 years, respectively. Time zero represents 1 year after ARNI initiation. Annualized event rates are expressed as events per 100 person-years. ARNI, angiotensin receptor-neprilysin inhibitor; GDMT, guideline-directed medical therapy; HF, heart failure.

Among patients with serial echocardiography, greater LV reverse remodeling was consistently associated with lower subsequent composite risk: adjusted HRs were 0.74 (95% CI, 0.67-0.80) per 5-percentage-point LVEF improvement, 0.82 (95% CI, 0.77-0.86) per 1% LVGLS improvement, 0.86 (95% CI, 0.82-0.90) per 10-mL LVEDV reduction, and 0.82 (95% CI, 0.78-0.86) per 10-mL LVESV reduction (all P<0.001) (**Supplementary Table S6**).

Exploratory indirect-effect analyses were subsequently performed to assess LV reverse remodeling as a potential intermediate pathway between GDMT intensity and the composite outcome (**Figure 4**). Bootstrap analyses with 1,000 resamples demonstrated significant indirect associations between greater GDMT intensity and lower subsequent composite risk through LVEF improvement (indirect HR, 0.92; 95% CI, 0.86-0.98), LVGLS improvement (indirect HR, 0.94; 95% CI, 0.88-0.99), LVEDV reduction (indirect HR, 0.93; 95% CI, 0.87-0.98), and LVESV reduction (indirect HR, 0.92; 95% CI, 0.85-0.98).

**Figure 4.**
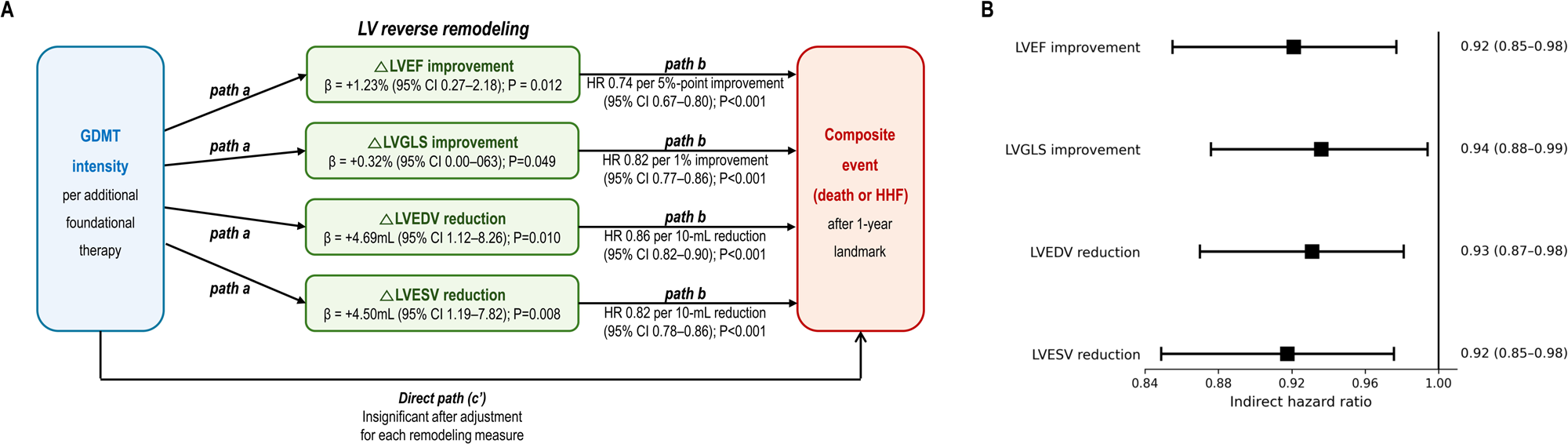
Exploratory Indirect-Effect Analysis of LV Reverse Remodeling. (A) LV reverse remodeling as a potential pathway between GDMT intensity and subsequent death or HF hospitalization. Remodeling measures were evaluated separately. (B) Indirect HRs with 95% CIs from 1,000 bootstrap resamples. CI, confidence interval; GDMT, guideline-directed medical therapy; HF, heart failure; HR, hazard ratio; LVEDV, left ventricular end-diastolic volume; LVEF, left ventricular ejection fraction; LVESV, left ventricular end-systolic volume; LVGLS, left ventricular global longitudinal strain.

## Discussion

In this multicenter ARNI-treated HFrEF cohort, comprehensive and four-pillar GDMT were less frequently achieved with advancing age, with renal dysfunction the most consistent correlate of incomplete treatment. Greater GDMT intensity was independently associated with more favorable LV reverse remodeling, which remained substantial at advanced age and strongly predicted subsequent prognosis. Although GDMT intensity itself was not independently associated with clinical outcomes, exploratory bootstrap analyses supported indirect associations through multiple LV remodeling measures. These findings suggest that myocardial recovery remains achievable at advanced age and may represent a pathway linking greater GDMT intensity with better prognosis.

Age-related underuse of evidence-based HF therapy has been consistently reported.^5–9^ Swedish and Korean registries demonstrated progressively lower use or achieved doses of recommended therapies with advancing age,^6,7^ and contemporary cohorts continue to show lower three-or four-pillar achievement in the oldest patients.^8,9^ Our findings extend this evidence to patients already receiving ARNI therapy: despite receipt of one foundational class, older patients remained less likely to receive additional pillars. Importantly, the clinical relevance of this treatment gap depends in part on whether older patients retain the capacity for treatment-associated myocardial recovery. Lower prescription intensity in older patients would be of less consequence if such capacity were substantially diminished; our findings suggest that meaningful recovery potential persists even at advanced age.

Treatment limitation in older patients is multifactorial. Renal dysfunction, hypotension, hyperkalemia, bradycardia, frailty, multimorbidity, and polypharmacy may influence treatment decisions.^5,6,9^ In our cohort, renal dysfunction was strongly associated with failure to achieve both comprehensive and quadruple GDMT. By contrast, diabetes was associated with greater implementation, possibly reflecting earlier SGLT2i use during a period when evidence, reimbursement, and clinical adoption of SGLT2i in HFrEF were evolving. Because observational associations cannot fully capture prior intolerance, treatment-related changes, or clinician selection, these findings should be interpreted as correlates rather than causal determinants of GDMT implementation.

The principal finding was the graded association between multidrug intensity and LV reverse remodeling. Each additional pillar was independently associated with greater improvement across complementary measures of LV functional and structural remodeling. Prior studies have demonstrated reverse remodeling with individual therapies, particularly sacubitril/valsartan.^14–18^ In PROVE-HF, LVEF and LV-volume improvement after sacubitril/valsartan was similar across age groups, supporting preserved remodeling potential at advanced age.^25^ A recent post-myocardial infarction cohort also reported a graded association between GDMT intensity and ventricular remodeling.^26^ Our study extends these observations by evaluating multidrug GDMT in HFrEF, demonstrating consistent associations across complementary measures of LV functional and structural recovery. Because treatment intensity was assessed as an ordinal exposure, these findings should not be interpreted as equivalent effects of individual classes; rather, they support a graded relationship between broader implementation of foundational therapy and the extent of reverse remodeling achieved during the first year of ARNI treatment.

The age-stratified findings should be interpreted cautiously. Although associations were strongest in patients aged <60 years, there was no significant age interaction for LVEF or LVGLS, and substantial recovery remained evident in older patients. Even among those aged ≥80 years, mean LVEF improved by approximately 10-12 percentage points with dual, triple, or quadruple therapy. The absence of significant trends in some older strata may reflect smaller sample sizes and limited quadruple-therapy use. Together with randomized-trial subgroup analyses showing preserved efficacy of ARNI, beta-blockers, and SGLT2i in older patients,^10–13^ these findings do not support withholding comprehensive GDMT solely because of advanced age when clinically tolerated. Rather, they suggest that advanced age does not eliminate the potential for clinically meaningful myocardial recovery with more intensive therapy.

The direct association between GDMT intensity and clinical outcomes was less definitive, with no independent association between pillar number and the composite endpoint. Such comparisons may be influenced by disease severity, renal function, frailty, and treatment tolerance. In contrast, achieved LV reverse remodeling was strongly associated with subsequent prognosis across LVEF, LVGLS, and LV volumes, consistent with prior STRATS-HF-ARNI studies.^15,17^ Exploratory indirect-effect analyses linked greater GDMT intensity with lower subsequent risk through LVEF, LVGLS, LVEDV, and LVESV changes, supporting myocardial recovery as one potential pathway connecting greater GDMT intensity with clinical outcomes. Nevertheless, these results should not be interpreted as proof of causal mediation. Because GDMT implementation and LV remodeling were assessed during the same first-year interval and treatment allocation was nonrandomized, temporal ordering and residual confounding remain important limitations. Thus, the indirect-effect analysis provides supportive evidence for a biologically and clinically coherent pathway rather than definitive evidence that reverse remodeling mediates the prognostic benefit of GDMT.

Finally, quadruple-therapy use increased markedly over calendar time, but age-related differences persisted overall. Importantly, older patients were those least likely to receive comprehensive GDMT despite retaining meaningful potential for LV recovery, and greater recovery was strongly associated with subsequent prognosis. Although treatment decisions in older patients require individualized consideration of renal function, hemodynamic tolerance, frailty, comorbidities, and patient preferences, advanced age itself should not be a reason to accept less comprehensive therapy. Comprehensive GDMT should therefore be actively pursued in older patients whenever clinically feasible and tolerated.

### Limitations

This study has several limitations. First, owing to the retrospective observational design, GDMT intensity was defined according to the number of drug classes rather than achieved doses, and detailed information regarding reasons for nonprescription, treatment intolerance, and dose modification during follow-up was not consistently available. In addition, GDMT intensity and reverse remodeling were assessed during the same first-year interval, limiting temporal inference and precluding causal interpretation. This temporal overlap is particularly relevant to the indirect-effect analysis because the temporal ordering required for formal causal mediation cannot be established; therefore, these findings should be interpreted as supporting a potential indirect pathway rather than demonstrating causal mediation. Second, the cohort was restricted to patients maintaining ARNI for ≥6 months, limiting generalizability to those unable to initiate or tolerate ARNI. Third, the overall achievement of four-pillar GDMT was relatively low, which may partly reflect the study period. Patients were enrolled between 2017 and 2022, during which evidence supporting ARNI and SGLT2i therapy in HFrEF was progressively established and incorporated into clinical guidelines. Moreover, reimbursement for ARNI and SGLT2i for HFrEF under the Korean National Health Insurance was introduced only in 2023 and 2024, respectively, which may have contributed to the relatively low use of four-pillar GDMT in this cohort. Fourth, follow-up echocardiography was nonuniform and less frequent in the oldest patients, which may have introduced selection bias in the reverse-remodeling analyses. Finally, prognostic analyses were exploratory and limited by small mono-and quadruple-therapy groups, particularly within age strata.

## Conclusions

Comprehensive and four-pillar GDMT were less frequently achieved with advancing age in ARNI-treated HFrEF. Greater GDMT intensity was independently associated with more favorable LV reverse remodeling, while substantial recovery remained possible at advanced age. Although GDMT intensity did not independently predict clinical events, greater achieved reverse remodeling was strongly associated with better prognosis. Exploratory bootstrap analyses further supported LV reverse remodeling as a potential intermediate pathway between greater GDMT intensity and lower subsequent clinical risk. These observational findings support efforts to reduce age-related treatment gaps and actively pursue comprehensive GDMT in older patients when clinically tolerated.

## Clinical Perspectives

### COMPETENCY IN MEDICAL KNOWLEDGE

Older patients with HFrEF remain less likely to receive comprehensive and four-pillar GDMT despite established ARNI therapy. Greater GDMT intensity is independently associated with more favorable LV reverse remodeling, with substantial myocardial recovery remaining evident even at advanced age.

### TRANSLATIONAL OUTLOOK

Prospective studies should evaluate strategies to safely increase comprehensive GDMT implementation in older patients and determine whether improved implementation is associated with greater myocardial recovery and, ultimately, better clinical outcomes.

## Abbreviations

ARNI: angiotensin receptor-neprilysin inhibitor
eGFR: estimated glomerular filtration rate
GDMT: guideline-directed medical therapy
HF: heart failure
HFrEF: heart failure with reduced ejection fraction
LV: left ventricular
LVEDV: left ventricular end-diastolic volume
LVEF: left ventricular ejection fraction
LVESV: left ventricular end-systolic volume
LVGLS: left ventricular global longitudinal strain

## Data Availability

The datasets used and/or analyzed during the current study are not publicly available because they contain clinical information subject to institutional and patient-privacy restrictions. The data may be available from the corresponding author upon reasonable request and subject to approval by the relevant institutional review boards.

## Acknowledgements

Not applicable.

## Authors’ contributions

ICH and HKK conceived and designed the study. ICH, JP, NYB, JL, SK, MB, HMC, JBP, YEY, SPL, YJK, GYC, and HKK contributed to data acquisition, analysis, or interpretation. ICH drafted the manuscript. All authors critically revised the manuscript for important intellectual content, approved the final version, and agreed to be accountable for the work.

## Central Illustration. Age-Related GDMT Implementation, Left Ventricular Reverse Remodeling, and Clinical Outcomes

ARNI, angiotensin receptor-neprilysin inhibitor; GDMT, guideline-directed medical therapy; HF, heart failure; HFrEF, heart failure with reduced ejection fraction; LV, left ventricular.

---

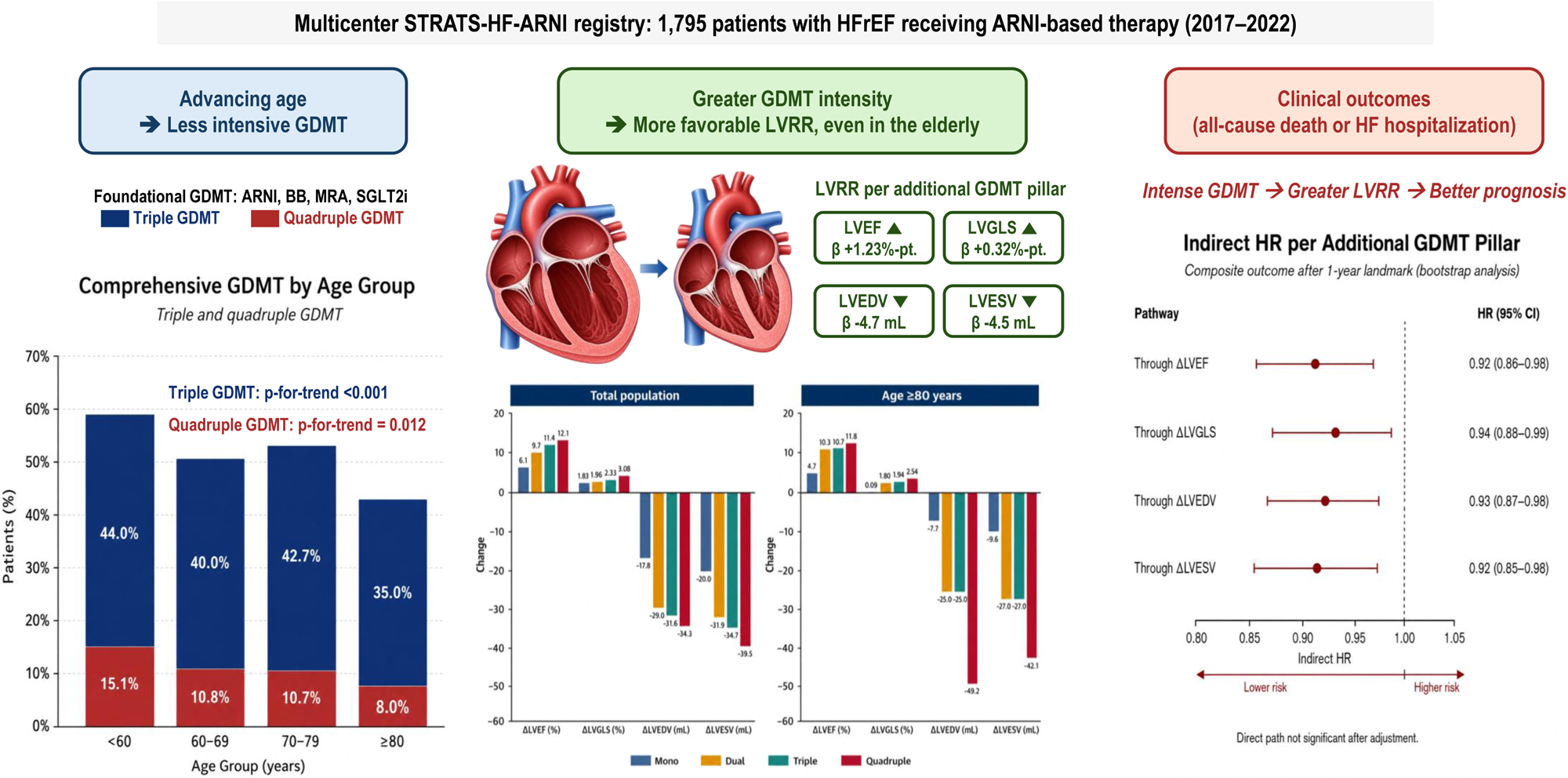

