## Supplementary materials for "Association of Age With Guideline-Directed Medical Therapy and Left Ventricular Reverse Remodeling"

**Supplementary Table S1. Actual Medication Combinations Among Patients Receiving Dual or Triple GDMT**

| **GDMT intensity** | **Actual combination** | **n** |
| --- | --- | --- |
| Dual | ARNI + BB | 672 (89.7%) |
|  | ARNI + MRA | 60 (8.0%) |
|  | ARNI + SGLT2i | 17 (2.3%) |
| Triple | ARNI + BB + MRA | 610 (82.4%) |
|  | ARNI + BB + SGLT2i | 121 (16.4%) |
|  | ARNI + MRA + SGLT2i | 9 (1.2%) |

Percentages are calculated within the corresponding GDMT-intensity group. Exact combination analyses included patients with complete BB, MRA, and SGLT2i medication data (dual therapy, n=749; triple therapy, n=740).

Abbreviations: ARNI, angiotensin receptor-neprilysin inhibitor; BB, beta-blocker; GDMT, guideline-directed medical therapy; MRA, mineralocorticoid receptor antagonist; SGLT2i, sodium-glucose cotransporter-2 inhibitor.

**Supplementary Table S2. Multivariable Factors Associated With GDMT Implementation in the Overall Population**

| **Variable** | **Failure to achieve triple/quadruple GDMT** | |  | **Failure to achieve quadruple GDMT** | |
| --- | --- | --- | --- | --- | --- |
|  | **Adjusted OR (95% CI)** | **P value** |  | **Adjusted OR (95% CI)** | **P value** |
| Age, per 10-year increase | 1.09 (1.01-1.18) | 0.036 |  | 1.19 (1.06-1.35) | 0.005 |
| Male | 1.13 (0.90-1.42) | 0.288 |  | 1.13 (0.79-1.63) | 0.498 |
| eGFR <60 mL/min/1.73 m² | 1.88 (1.49-2.36) | <0.001 |  | 2.26 (1.53-3.33) | <0.001 |
| Diabetes mellitus | 0.51 (0.40-0.64) | <0.001 |  | 0.17 (0.12-0.23) | <0.001 |
| Atrial fibrillation | 0.92 (0.73-1.17) | 0.504 |  | 0.94 (0.64-1.39) | 0.770 |
| Coronary artery disease | 1.27 (1.01-1.58) | 0.037 |  | 1.17 (0.82-1.67) | 0.396 |
| Baseline LVEF, per 10% increase | 1.30 (1.14-1.48) | <0.001 |  | 1.47 (1.19-1.82) | <0.001 |
| Calendar year, per 1-year increase | 0.88 (0.82-0.94) | <0.001 |  | 0.55 (0.48-0.63) | <0.001 |

Both multivariable models included 1,593 patients with complete data for the included covariates. An OR >1 indicates a higher likelihood of failure to achieve the corresponding GDMT threshold (triple/quadruple GDMT or quadruple GDMT).

Abbreviations: CI, confidence interval; eGFR, estimated glomerular filtration rate; GDMT, guideline-directed medical therapy; LVEF, left ventricular ejection fraction; OR, odds ratio.

**Supplementary Table S3. Multivariable Factors Associated With Failure to Achieve Triple/Quadruple GDMT in Patients Aged ≥70 Years**

| **Variable** | **Adjusted OR (95% CI)** | **P value** |
| --- | --- | --- |
| Age, per 10-year increase | 1.47 (1.08-2.02) | 0.016 |
| Male | 1.31 (0.95-1.81) | 0.100 |
| eGFR <60 mL/min/1.73 m² | 1.80 (1.30-2.50) | <0.001 |
| Diabetes mellitus | 0.50 (0.35-0.71) | <0.001 |
| Atrial fibrillation | 0.87 (0.62-1.23) | 0.428 |
| Coronary artery disease | 1.19 (0.85-1.67) | 0.307 |
| Baseline LVEF, per 10% increase | 1.22 (1.00-1.49) | 0.055 |
| Calendar year, per 1-year increase | 0.82 (0.74-0.91) | <0.001 |

The multivariable analysis included 681 patients aged ≥70 years with complete data for the included covariates. An OR >1 indicates a higher likelihood of failure to achieve triple/quadruple GDMT.

Abbreviations: CI, confidence interval; eGFR, estimated glomerular filtration rate; GDMT, guideline-directed medical therapy; LVEF, left ventricular ejection fraction; OR, odds ratio.

**Supplementary Table S4. Association of GDMT Intensity With Clinical Outcomes in 1-Year Landmark Analyses**

| **Outcome** | **GDMT comparison** | **Adjusted HR (95% CI)** | **P value** |
| --- | --- | --- | --- |
| **All-cause death** | Mono | Reference |  |
|  | Dual | 0.41 (0.19–0.90) | 0.026 |
|  | Triple | 0.48 (0.22–1.06) | 0.070 |
|  | Quadruple | 0.25 (0.06–0.99) | 0.049 |
| **HF hospitalization** | Mono | Reference |  |
|  | Dual | 0.93 (0.40–2.17) | 0.871 |
|  | Triple | 1.07 (0.46–2.50) | 0.867 |
|  | Quadruple | 0.99 (0.35–2.77) | 0.987 |
| **Composite of all-cause death and HF hospitalization** | Mono | Reference |  |
|  | Dual | 0.76 (0.39–1.48) | 0.425 |
|  | Triple | 0.81 (0.41–1.57) | 0.524 |
|  | Quadruple | 0.67 (0.28–1.60) | 0.368 |

All analyses were performed using a 1-year landmark approach. Patients who experienced the corresponding outcome within 1 year after ARNI initiation or were no longer under observation at the landmark were excluded from the respective analysis.

Multivariable models were adjusted for age, sex, participating center, systolic blood pressure, estimated glomerular filtration rate, serum potassium, diabetes mellitus, atrial fibrillation, coronary artery disease, baseline left ventricular ejection fraction, and calendar year of ARNI initiation.

Abbreviations: ARNI, angiotensin receptor–neprilysin inhibitor; CI, confidence interval; GDMT, guideline-directed medical therapy; HF, heart failure; HFH, heart failure hospitalization; HR, hazard ratio.

**Supplementary Table S5. Multivariable Predictors of the Composite of All-Cause Death or Heart Failure Hospitalization in the 1-Year Landmark Analysis**

| **Variable** | **Adjusted HR (95% CI)** | **P value** |
| --- | --- | --- |
| Age, per 10 years | 1.37 (1.21–1.56) | <0.001 |
| Male sex | 1.25 (0.91–1.71) | 0.171 |
| SBP, per 10 mmHg | 0.93 (0.87–1.01) | 0.071 |
| eGFR, per 10 mL/min/1.73m² | 0.87 (0.82–0.93) | <0.001 |
| Diabetes mellitus | 0.91 (0.66–1.26) | 0.572 |
| Atrial fibrillation | 1.22 (0.89–1.66) | 0.211 |
| Coronary artery disease | 1.03 (0.76–1.39) | 0.870 |
| Baseline LVEF, per 10% | 0.84 (0.70–1.02) | 0.074 |
| GDMT intensity, per additional pillar | 0.94 (0.76–1.16) | 0.545 |

The multivariable model included 1,058 patients with complete covariate data and 199 events and was adjusted for all variables shown.

Abbreviations: CI, confidence interval; eGFR, estimated glomerular filtration rate; GDMT, guideline-directed medical therapy; HR, hazard ratio; LVEF, left ventricular ejection fraction; SBP, systolic blood pressure.

**Supplementary Table S6. Associations of Echocardiographic Reverse Remodeling With the Composite Clinical Outcome**

| **Echocardiographic change** | **Adjusted HR (95% CI)** | **P value** |
| --- | --- | --- |
| ΔLVEF, per +5% improvement | 0.74 (0.67–0.80) | <0.001 |
| ΔLVGLS, per +1% improvement | 0.82 (0.77–0.86) | <0.001 |
| LVEDV reduction, per 10 mL | 0.86 (0.82–0.90) | <0.001 |
| LVESV reduction, per 10 mL | 0.82 (0.78–0.86) | <0.001 |

Each echocardiographic change was evaluated in a separate 1-year landmark Cox model because of collinearity among remodeling measures. Models were adjusted for relevant clinical covariates and the corresponding baseline echocardiographic parameter. Favorable changes were defined as increases in LVEF and LVGLS, and reductions in LVEDV and LVESV.

Abbreviations: CI, confidence interval; HR, hazard ratio; LVEDV, left ventricular end-diastolic volume; LVEF, left ventricular ejection fraction; LVESV, left ventricular end-systolic volume; LVGLS, left ventricular global longitudinal strain.

**Supplementary Figure S1. Clinical Outcomes According to Age Group**

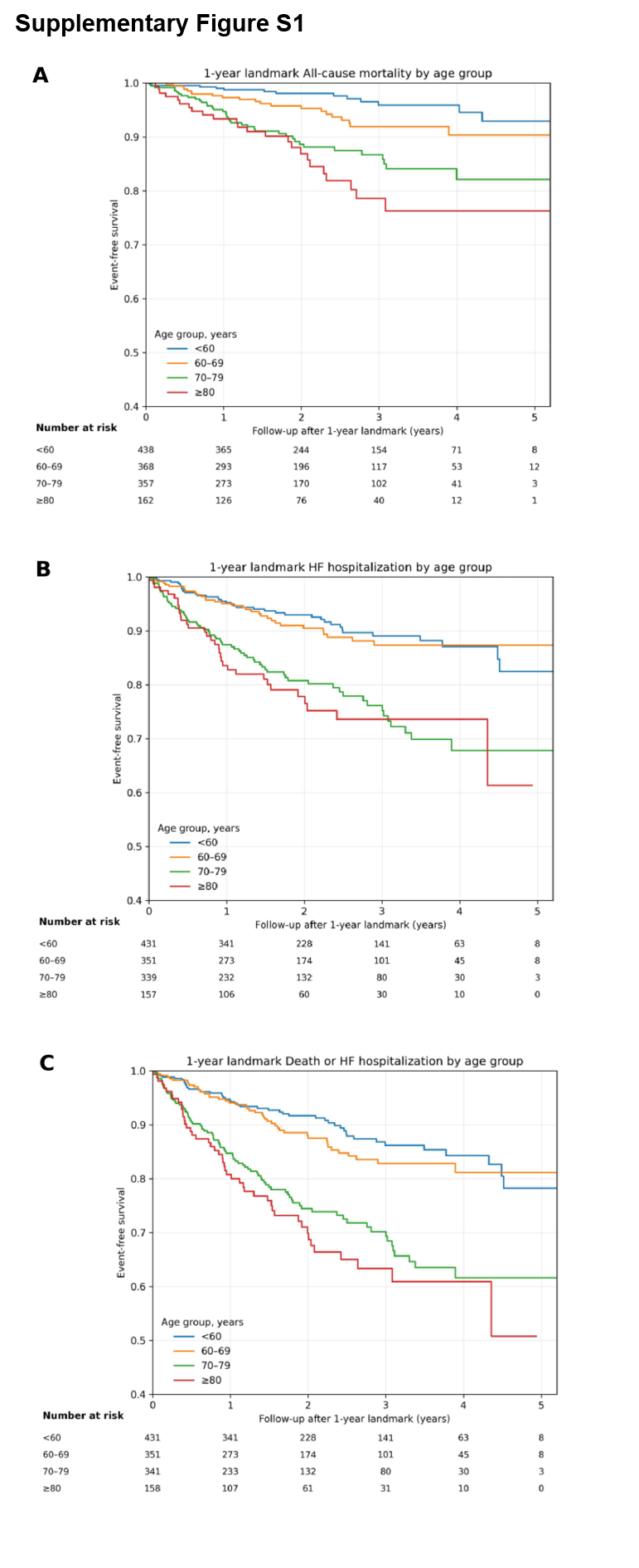

One-year landmark Kaplan-Meier curves for (A) all-cause mortality, (B) HF hospitalization, and (C) their composite according to age. Time zero represents 1 year after ARNI initiation.

ARNI, angiotensin receptor-neprilysin inhibitor; HF, heart failure.

**Supplementary Figure S2. All-Cause Mortality According to GDMT Intensity and Age in the 1-Year Landmark Analysis**

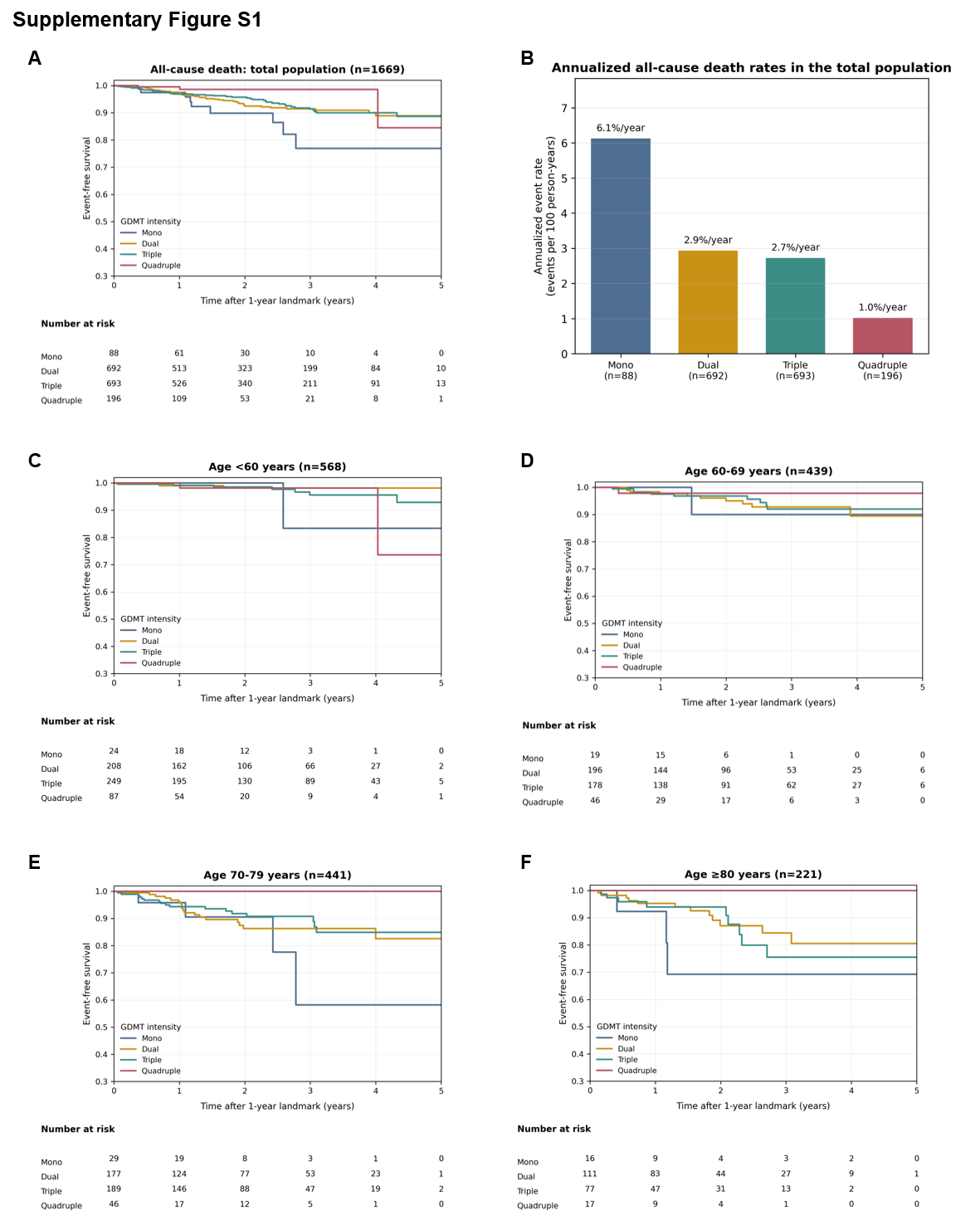

One-year landmark analysis of all-cause mortality according to mono-, dual-, triple-, and quadruple GDMT. (A) Kaplan-Meier curves in the total population. (B) Annualized all-cause mortality rates according to GDMT intensity in the total population. (C–F) Kaplan-Meier curves in patients aged <60, 60–69, 70–79, and ≥80 years, respectively. Annualized event rates are expressed as events per 100 person-years. GDMT, guideline-directed medical therapy.

**Supplementary Figure S3. Heart Failure Hospitalization According to GDMT Intensity and Age in the 1-Year Landmark Analysis**

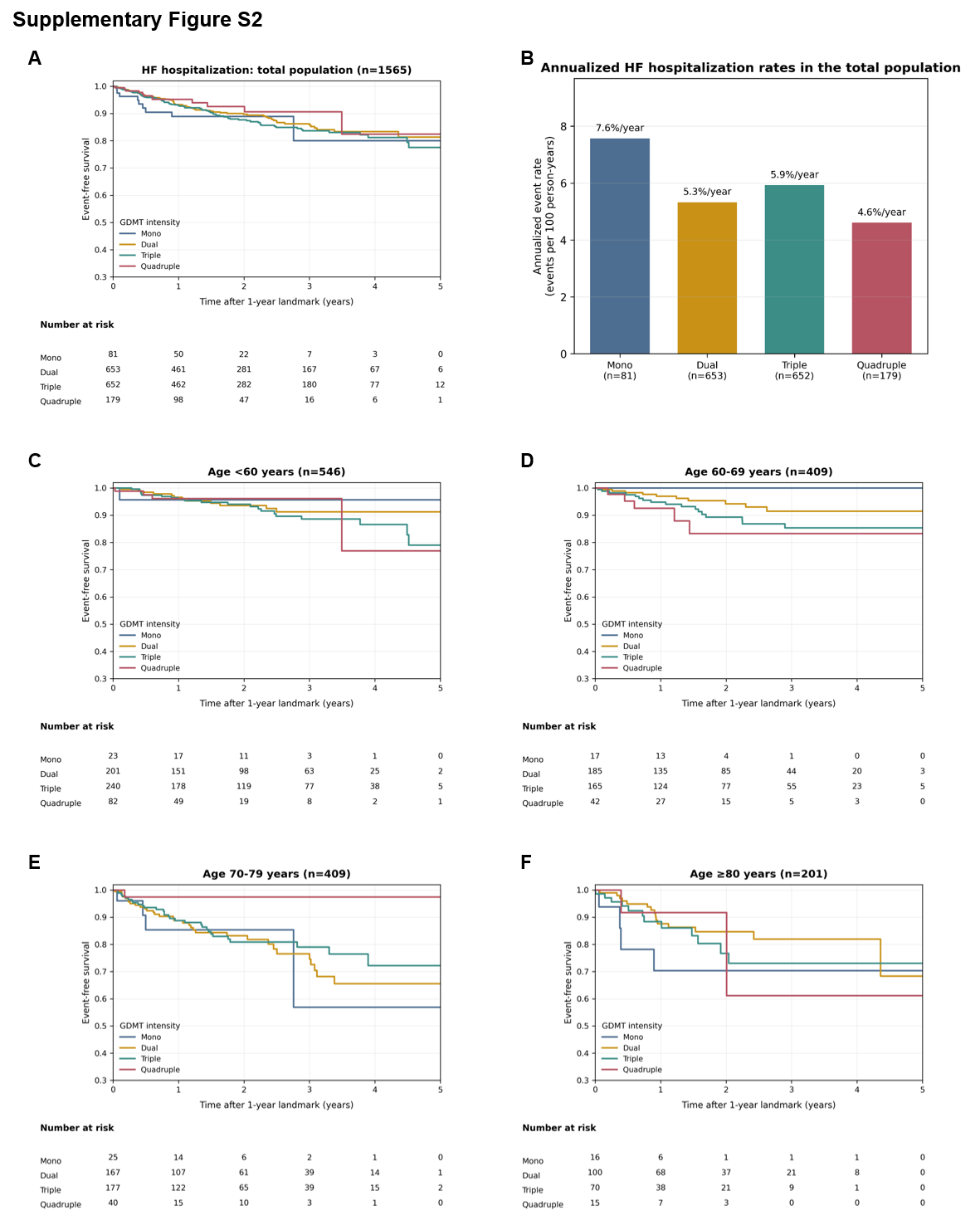

One-year landmark analysis of HF hospitalization according to mono-, dual-, triple-, and quadruple GDMT. (A) Kaplan-Meier curves in the total population. (B) Annualized HF hospitalization rates according to GDMT intensity in the total population. (C–F) Kaplan-Meier curves in patients aged <60, 60–69, 70–79, and ≥80 years, respectively. Annualized event rates are expressed as events per 100 person-years. GDMT, guideline-directed medical therapy; HF, heart failure.
